# Remote Digital Cognitive Assessment Reveals Cognitive Deficits Related to Hippocampal Atrophy in Autoimmune Limbic Encephalitis

**DOI:** 10.1101/2023.07.25.23292765

**Authors:** Kengo Shibata, Bahaaeddin Attaallah, Xin-You Tai, William Trender, Peter J. Hellyer, Adam Hampshire, Sarosh R Irani, Sanjay G Manohar, Masud Husain

**Affiliations:** Nuffield Department of Clinical Neurosciences, University of Oxford, Oxford OX3 9DU, UK; Department of Brain Sciences, Imperial College London, London, W12 0NN UK; Department of Experimental Psychology, University of Oxford, Oxford OX1 3PH, UK; Centre for Neuroimaging Sciences, King’s College London, London SE5 8AF

**Keywords:** <u>Keywords</u>: autoimmune limbic encephalitis, hippocampus, remote digital cognitive testing, online cognitive assessment, cognitive profile

## Abstract

Autoimmune Limbic Encephalitis (ALE) is a neurological disease characterised by inflammation of the limbic regions of the brain, mediated by pathogenic autoantibodies. Because cognitive deficits persist following acute treatment of ALE, the accurate assessment of long-term cognitive outcomes is important for clinical assessments and trials. However, evaluating cognition is costly and an unmet need for validated digital methods exists. We investigated whether remote digital methods could identify previously characterised cognitive impairments in ALE patients and would correlate with standard neuropsychological assessment and hippocampal volume. The cognitive performance of 21 chronic ALE patients along with 54 age-matched healthy controls was assessed with a battery of 12 cognitive tasks from the Cognitron online platform. ALE patients performed significantly worse in memory, visuospatial abilities, executive function, and language. No impairments in digit & spatial span, target detection (attention) and emotion discrimination were observed. The global score on the online cognitive tasks correlated significantly with the established Addenbrooke’s Cognitive Examination III (ACE) pen-and-paper test. Deficits in visuospatial processing and language were identified in ALE compared to controls using remote digital testing but not the ACE, highlighting higher sensitivity of computerised testing to residual cognitive impairment. Finally, the hippocampal volume of ALE patients and healthy controls correlated with online cognitive scores. Overall, these findings demonstrate that remote, online testing may facilitate the characterisation of cognitive profiles in complex neurological diseases.

## Introduction

Autoimmune limbic encephalitis (ALE) is an inflammatory disease that affects the structural integrity and functioning of the limbic system. Two frequently presented sub-types of ALE are characterised by autoantibodies against Leucine-rich glioma-inactivated 1 (LGI1) and Contactin-associated protein 2-Antibody (CASPR2)^1^. These antibodies are considered pathogenic and their transfer to rodents recapitulates cognitive aspects of the human conditions^2, 3^. Clinically, ALE patients present with seizures, deficits in cognition and neuropsychiatric symptoms^4–6^. However, the specific clinical and neuroimaging manifestations in these forms of ALE can vary widely across patients^4, 5, 7, 8^. Despite being a common feature, long-term cognitive deficits caused by ALE are still not fully characterised^9^. Remote, multi-dimensional quantitation of patients’ cognitive profiles could help track disease activity and response to immunotherapies and are especially timely given commencement of clinical trials in this field.

Immunotherapies improve outcomes in several ALE syndromes^4, 7, 10, 11^ with most patients showing major improvements on the modified Rankin Scale (mRS) measure of disability and dependence on daily activities. Although cognition often improves on bedside testing (Montreal Cognitive Assessment (MoCA) and the Mini-Mental State Examination (MMSE)^12, 13^, residual and persistent deficits in memory and executive function are common^14^ even several years post-treatment^4, 9, 12, 15–17^. Specifically, for LGI1-antibody encephalitis patients, deficits in episodic memory^7, 18^, working memory^18, 19^, language^12^ and fluency^12^ as well as fatigue^9^ are reported in post-acute phases of the disease. Despite this, long-term cognitive outcomes are not routinely assessed, likely due to the resources required to carry out in-person testing and the lack of validated tools.

Previous studies have found that surrogate digital measures may provide a rapid and cost-effective means of measuring long-term cognitive outcomes in a range of neurodegenerative disorders such as mild cognitive impairment (MCI)^20^, and Alzheimer’s disease (AD)^21^. Recently, self-directed web-based computerised tasks have shown high validity with established clinical tests for MCI^22^ and sensitive to changes in traumatic brain injuries (TBI)^23^ and early AD^24^. Computerised testing also provides the opportunity to analyse trial-by-trial data to dissociate cognitive abilities from visuomotor response latencies^25^. Such approaches highlight the use case of high-dimensional data that computerised testing can offer which cannot be generalised to pen-and-paper testing. Although further research is needed to validate the use of digital measures in clinical settings, these studies highlight the use of this technology to improve cognitive testing and overcome some of the limitations of in-person clinical assessments. Finally, the COVID-19 pandemic led to an increased demand for remote, contactless cognitive assessments. Investigations during the pandemic^26–28^ have demonstrated the feasibility of home digital testing across different age groups.

In this study, we use the Cognitron online cognitive assessment tool to investigate the cognitive profile of chronic LGI1-and CASPR2-antibody encephalitis patients; two subtypes of ALE with similar neuropsychological profiles^29^. Our objective was to identify domain-specific cognitive deficits using digital cognitive testing and correlate the results with the core behavioural and neuropsychiatric features of ALE. We compared performance on this digital testing platform with Addenbrooke’s Cognitive Examination - III (ACE), a standard in-person clinical screening examination^30^. In light of previous reports showing hippocampal atrophy in ALE patients and associated cognitive deficits^1, 5, 7^, we also assessed the relationship between hippocampal volume and cognitive outcomes. The evaluation of online cognitive assessment tools has the potential to facilitate the in-depth characterisation of long-term cognitive outcomes in ALE with important clinical utility.

## Materials & Methods

### Participants

Twenty-one chronic ALE patients (mean age = 63.01, SD = 8.14, 14 males) and 54 healthy controls (mean age = 65.56, SD = 7.31, 23 males) enrolled in the University of Oxford’s Cognitive Neurology Lab testing program. All ALE patients had been clinically assessed and diagnosed by neurologists (M.H., S.M. and S.I.) at John Radcliffe Hospital in Oxford, UK. Cognitive assessment using Addenbrooke’s Cognitive Examination III (ACE; Hsieh et al., 2013) was performed in 50/54 healthy controls and 20/21 ALE patients. Magnetic resonance imaging was acquired in 47/54 healthy controls and 21/21 ALE patients. Participants provided written informed consent and were offered monetary compensation for their participation. This study was approved by the University of Oxford’s Ethics Committee (18/SC/0048 & REC16/YH/0013).

### Behavioural Paradigms

ALE patients and healthy controls were assessed using a modified version of the Cognitron protocol, an online battery of cognitive tasks that run via web browsers (available at: https://ompilot2.cognitron.co.uk/). The battery of tasks was programmed in HTML5 with JavaScript and hosted on the Amazon EC2 platform. The platform was optimised with National Institute for Health and Care Research (NIHR) support to be deliverable remotely to elderly and patient groups. This battery comprised 12 short cognitive tasks run in the same order for all participants, which are summarised below. Participants accessed the tasks through a link on a web browser and were asked to run them on their own computers. Written instructions were provided at the start of the assessment and then at the start of each task. Feedback was provided after the completion of all tasks. The average delay time between the immediate object memory task (run first) and the delayed object memory task (run last) was comparable between ALE patients (42.45 minutes, SD: 7.76) and healthy controls (39.13 minutes, SD: 5.99), (t(29.76) = -1.77, p = .087).

### 1. Object Memory

Twenty black-and-white images of everyday objects were presented sequentially in random order. Presentation times were uniform at 2000 ms, with an inter-stimulus interval of 500 ms. Participants were tested on their memory of the objects immediately after the presentation in a multiple-choice array of 8 images (a 3 by 3 grid with an empty central box, Figure 2A). One image was identical to one of the 20 objects presented prior and was the correct answer. The multiple-choice array also consisted of the identical but mirrored image, an object from the same semantic category (and its mirrored version) and 4 other images from a separate semantic category. Errors were categorised as either: 1) Spatial Error, 2) Item Error or 3) Category Error (guess), where Correct + Spatial Error + Item Error + Category Error = 1 (Figure 2B). Delayed recall of the objects was tested a second time after all other cognitive tasks were completed. Participants were not informed that the memory of these objects would be probed at a delayed time point. The proportion of correct answers was used as the primary metric for object memory.

**Figure 1.**
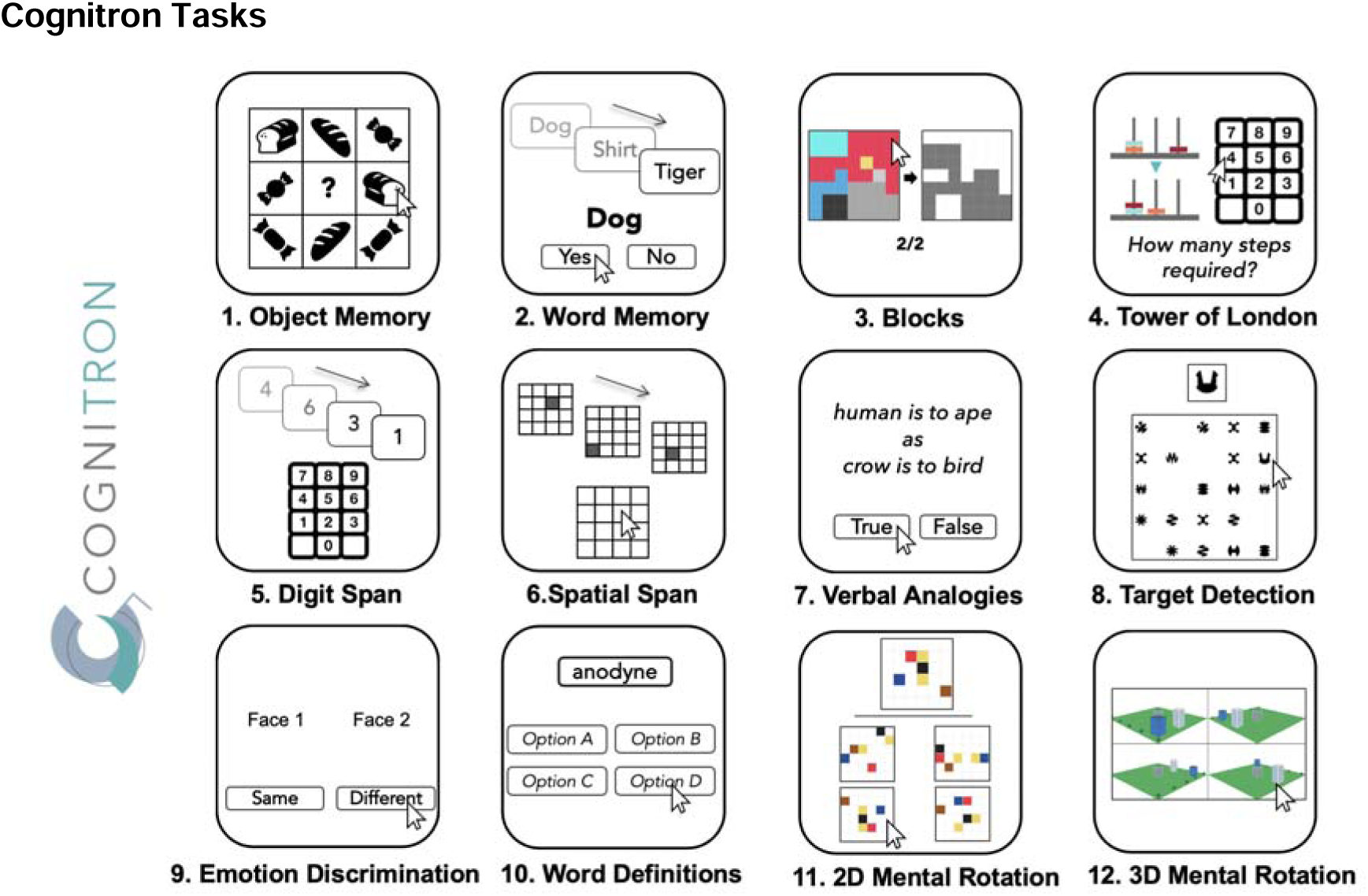
Online Cognitron battery. Twelve different cognitive tasks make up the Cognitron battery. Tasks were administered in the presented order, with Object Memory and Word Memory being tested at a delayed time point after all other tasks were completed. Feedback was provided after all tasks. The tasks were all run remotely in one sitting.

**Figure 2.**
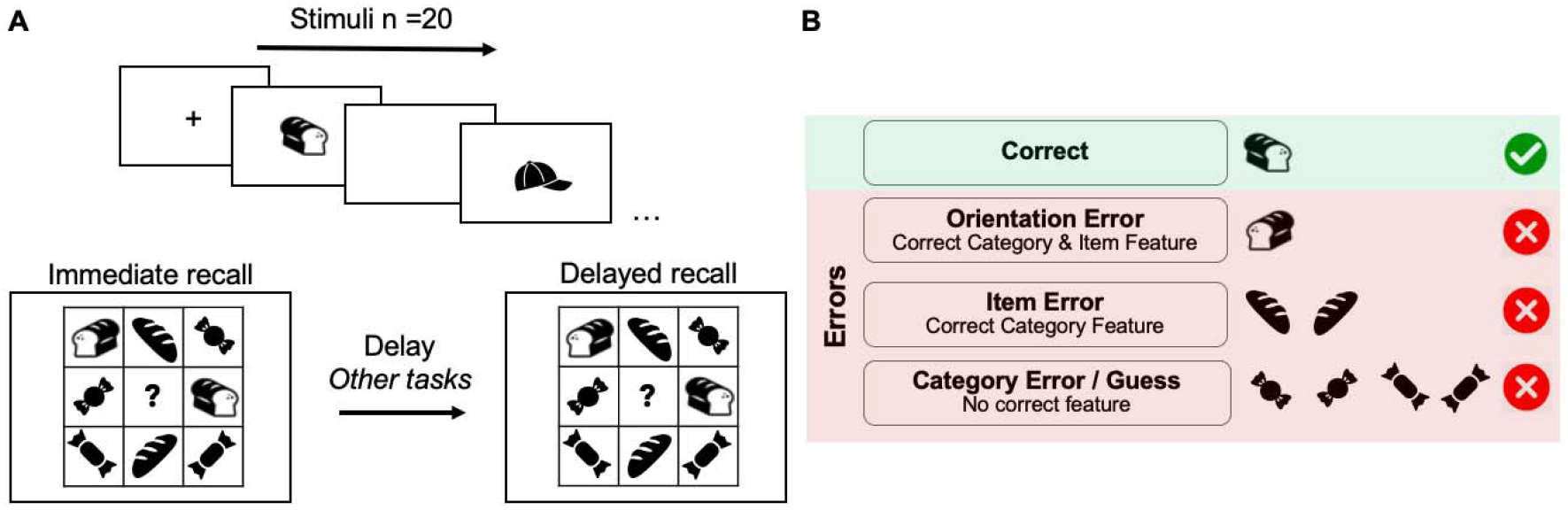
Object Memory task. A) Twenty objects are presented sequentially. Immediately after the presentation, participants were required to identify an object presented in the initial sequence within an array of 8 objects of varying similarities, in random order. The same recognition task was administered at the end in delayed recall, on average 40 minutes after. B) Task performance was calculated using accuracy of correct answers. The errors made were categorised into three different precision measures of recognition. Each has a varying number of correct features, in descending order.

### 2. Word Memory

Twelve English words were presented sequentially for 1000 ms with an interstimulus interval of 200 ms. The words were drawn from three categories: animals, clothes, and vegetables. Participants were asked to categorise a list of 24 words into old (seen in the earlier sequence) or new; 50% were targets (seen in the initial sequence of 12 words), and 50% were non-targets, of which half were lures with semantic similarity to the targets. Participants were also tested on the same set of words in a delayed recall after all other tasks in the battery, just after the delayed recall for the Object Memory Task. The delay time was comparable to that of the Object Memory delay. Performance correct, correct rejection of foil and lures were calculated. The number of correct answers was used as the primary performance metric.

### 3. Blocks

To assess spatial planning ability, participants were asked to match a configuration of tiles by removing shapes from a pre-configured layout of tiles. Importantly, the unsupported tiles fell down with gravity, changing the configuration. No time limit was enforced. The number of correct trials was used as the outcome variable.

### 4. Tower of London

A modified version of the Tower of London task was used to investigate spatial planning abilities. Participants were presented with two sets of three prongs with coloured beads on them. They were required to count the minimum number of moves it would take to change one set of three prongs to the other configuration and indicate the number on a number pad. Participants could not directly move the coloured beads and had to hold the number of moves in memory. There were 10 trials of varying difficulty, and the proportion correct was used as a measure of mental spatial planning.

### 5. Digit span

The digit span task required participants to hold numbers shown sequentially in memory. On each trial, participants were presented with a series of digits. At the end of the sequence, participants were required to recall the digits in the order they appeared by clicking on a number pad. Each successful trial led to an increase in the number of digits by one. The task was terminated when three consecutive errors were made on a given difficulty level. The maximum attained span was used as the memory measure, which ranged from 1 to 20.

### 6. Spatial Span

Spatial short-term memory was tested using an adapted version of the Corsi Block Tapping Test. A sequence of locations was lit up in a 4 by 4 grid, and participants had to replicate the sequence. All trials began with two grids lighting up sequentially and were incremented by one on every successful recall. The task was terminated when three consecutive errors were made on a given difficulty level. The maximum sequence length for each participant was taken as a measure of spatial memory span, which ranged from 1 to 16.

### 7. Verbal Analogies

Semantic (analogical) reasoning was tested by assessing the associations of two pairs of words. Participants were asked to indicate whether the association between words was true or false (e.g., ‘Human is to ape as crow is to bird’, true). Participants answered as many prompts as possible in 3 minutes. Every incorrect response led to a deduction of one point. Every correct point led to an increase of one point. The total score was used as a measure of word-based semantic reasoning.

### 8. Target Detection

Spatio-visual attention was tested by presenting a target shape amongst a grid of dynamically changing shapes. The target shape remained on the left side of the screen for reference and remained unchanged for a given participant. In a 5 by 5 grid, new shapes were added every 1s and others removed every 1s. A target shape was added pseudo-randomly, at a frequency of 12 in every 20 new shapes. Participants were instructed to click on as many target shapes as possible. The task ended after 120 addition/removal cycles. The total number of target shapes clicked on was used as a measure of visuospatial target detection.

### 9. Emotion Discrimination

The emotion discrimination task tested how well emotions are recognized by discriminating whether two facial emotions of different identities were the same or different. 50 trials were presented sequentially, and one point was awarded for every correct answer. No time limit was enforced in this task. The proportion of correct responses was used as the measure of emotion discrimination.

### 10. Word Definitions

The word definition task tested semantic ability. Under each word, four definitions were provided. Each trial had to be completed within 20 seconds, and participants had to click on one of the four choices that most accurately defined the word. The proportion correct out of 21 trials was calculated.

## 11. 2D Mental Rotation

2D mental rotation was tested by presenting a 6 by 6 grid with a specific arrangement of coloured cells. This was the target grid presented next to four other grids. One of the four grids was an identical but rotated version of the target grid, which had to be selected. Participants had 3 minutes to complete as many trials as possible. The proportion of correct trials was used as a measure of 2D mental rotation.

## 12. 3D Mental Rotation

3D mental rotation was assessed by having participants find the rotated version of a 3D image of buildings that was the odd one out, where three configurations were identical when rotated. The task consisted of 12 trials, and the proportion of correct choices was used as a measure of 3D mental rotation.

### Magnetic Resonance Data Acquisition

Structural Magnetic Resonance Images (MRI) were obtained from 20/21 ALE patients and 47/54 healthy controls who were MRI-compatible. Participants were scanned at the John Radcliffe Hospital in Oxford using a 3T Siemens Verio scanner. T1-weighted structural images had 1mm isotropic voxel resolution (MPRAGE, field of view: 208×256×256 matrix, TR/TE = 200/1.94 ms, flip angle = 8 ∘ 618, iPAT =2, pre-scan-normalise).

### Magnetic Resonance Data processing and Analysis

Images were pre-processed with FMRIB Software Library (FSL) according to the UK Biobank analysis pipeline (https://git.fmrib.ox.ac.uk/falmagro/UK_biobank_pipeline_ v_1)^31^. Hippocampal volumes were extracted from T1 anatomical images using FSL FIRST for segmentation and FSL SIENAX for volume extraction. Brain volumes were corrected for head size by the following equation^32^:

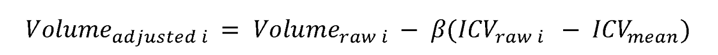

Where β is the slope of the line of regression between the ICV (Intracranial volume) and the adjusted volume.

### Statistical Analysis

Statistical analysis was carried out on MATLAB R2021b and R Version 1.3.959. Equality of variance was tested using Levene’s test and two-tailed Welch’s t-test, assuming unequal variance between groups, was used for group comparison. Linear regressions were run by correcting for gender, age and years of education, and visualised using the *plotAdded* function on MATLAB. Corresponding p and r values are reported. To provide a global score for cognition, a principal component analysis of the main outcome measure of all tasks was performed using the *pca* function in MATLAB. The first principal component was extracted as a global measure of cognition. The same dimensionality reduction analysis was carried out for the clustered domains of the Cognitron task set that shared cognitive processes. Network plots for inter-task correlations were assessed and visualised using the *network_plot* function in R, where a stronger correlation between tasks is represented by their spatial proximity and the colour of the lines connecting them. Mixed effect modelling was performed using the *fitglm* function in MATLAB.

## Results

### Demographics

Healthy controls and ALE patients did not differ in gender or in age. ALE patients had fewer years of education and, as expected, a lower total ACE score (Table 1). Out of the 21 ALE patients, 15 had LGI1-antibodies, and 6 had CASPR2-antibodies.

**Table 1.** Study demographics. Healthy controls do not differ overall in age or gender to autoimmune limbic encephalitis (ALE) patients. The average years of education are higher in the healthy controls, although a high average of 12.45 years is reported for patients.

|  | All<br>(n = 75) | Healthy control<br>(n = 54) | ALE<br>(n = 21) | Statistics |
| --- | --- | --- | --- | --- |
| Gender<br>Female (%) | 38 (50.67) | 31 (57.41) | 7 (33.33) | $\chi^2(1, N = 75) = 3.51, p = .06$ |
| Age<br>Mean years (SD) | 64.89 (7.56) | 65.56 (7.31) | 63.19 (8.10) | $t(33.35) = 1.16, p = .25, d = 0.31$ |
| Years of Education<br>Mean years (SD) | 14.41 (2.97) | 15.23 (2.81) | 12.45 (2.44) | **$t(40.76) = 4.09, p = .0002, d = 1.03$** |
| ACE-III<br>Mean score (SD) | 96.03 (4.20) | 97.46 (1.89) | 92.45 (6.00) | **$t(20.52) = 3.67, p = .0015, d = 1.42$** |

### Comparison of ALE and healthy controls

The average group performance was assessed for each task. Healthy controls outperformed ALE patients in 10 out of 14 tasks, (p < .05, d > 0.7). (Figure 3A, See Supplemental Table 1 for statistics of group comparisons). In the neuropsychological ACE, healthy controls outperformed ALE patients in the memory and fluency domains, but no difference was observed for language, visuospatial and attention domains (Figure 3B, See Supplemental Table 2 for details of group comparisons).

**Figure 3.**
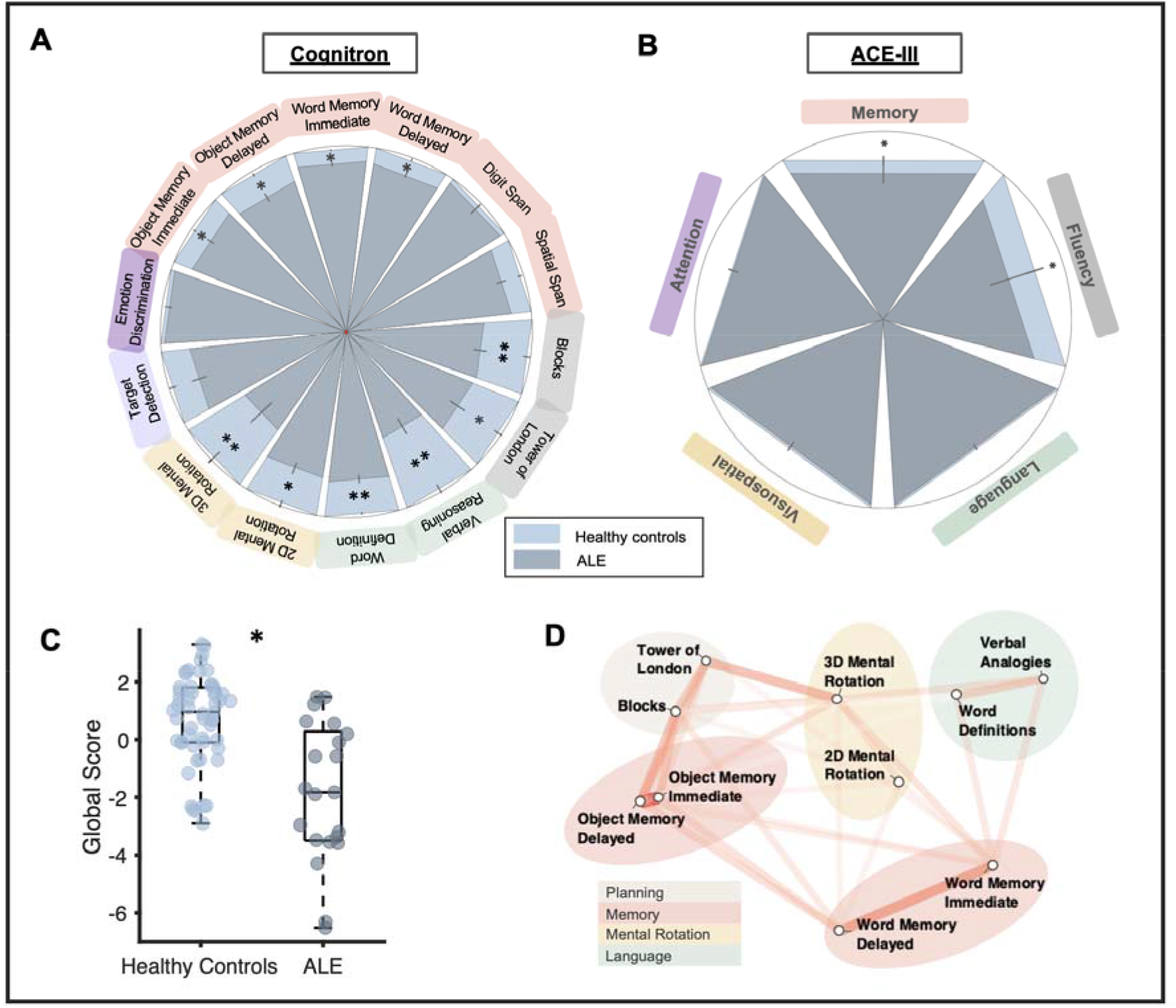
Performance on Cognitron and ACE. A) Group differences in performance on each task were tested using Welch’s t-test and visualised with a circular plot. Performance is scaled to the baseline performance of healthy controls (in light blue). Bars closer to the centre represent lower performance. * = <.05, ** = <.01. Errors bars are SEM. B) Group difference in ACE domains also using Welch’s t-test revealed a difference in memory and fluency, but not in language, visuospatial abilities, or attention. Performance is scaled to the baseline performance of healthy controls. C) A lower Global Score, calculated as the first principal component of all cognitive tasks combined, was found in ALE patients compared to healthy controls. D) Network plot of task correlations (healthy controls only) reveals task clustering. Only metrics that significantly correlated with another task are included in the plot. A smaller distance and lower opacity of the connecting line indicate a stronger correlation.

### Clustering of online tasks yields five traditional cognitive domains

The first principal component from all cognitive measures of the computerised tasks was taken as a global cognition score. This component explained 32% of the variance in the dataset. The global score of cognition was higher in healthy controls (M = 0.71, SD = 1.53) compared to ALE patients (M = -1.83, SD = 2.40), t(20.52) = 3.67, p = .0015, d = 1.42 (Figure 3C). Analysis of correlations between individual task performances on healthy controls showed that the Cognitron tasks could be clustered into five domains: memory, planning, mental rotation, and language (Figure 3D). Similar groupings were obtained with the patient group (Supplemental Figure 1). The strongest correlation observed in the Cognitron tasks was between the immediate and delayed version of the object and word memory tasks (object: r(73) = 0.77, p <.001; word r(73) = 0.59, p <.001).

### Cognitron Performance Correlates with ACE scores

We investigated the relationship between the global Cognitron score and ACE scores as a validation metric. Across all participants, the global Cognitron score was highly correlated with total ACE scores, whilst controlling for age, gender and years of education (r(49) = 0.46, p < .001, Figure 4). This correlation remained significant when tested in ALE patients separately (r(19) = 0.57, p = .02) but not in healthy controls (r(49) = 0.09, p = .09,). This relationship is likely to be constrained by the limited variance of the ACE score in the healthy controls.

**Figure 4.**
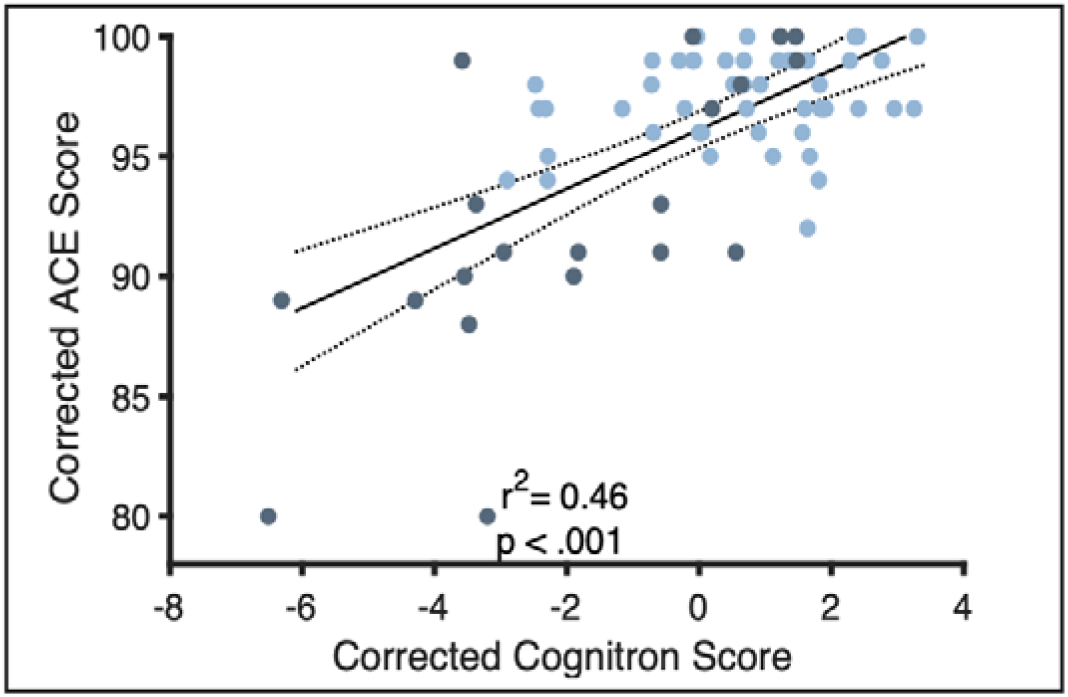
Correlation between clinical ACE score and digital Cognitron score. Linear regression between ACE score and Cognitron scores. The regression line is corrected for age, years of education and gender. The lighter blue dots indicate healthy controls, and the darker dots indicate ALE patients.

We took the first principal component of all computerised tasks in a single cognitive domain to compose a subdomain composite score (as grouped in the network plot, Figure 3C). Each subdomain from the Cognitron battery was correlated with its corresponding ACE subdomain (memory, language, verbal fluency, visuospatial abilities, and attention). There was a significant correlation between Cognitron scores and the in-lab neuropsychological assessments in memory, attention and language domains (Supplemental Figure 2) showing that Cognitron domains mapped onto clinical neuropsychological test scores. Furthermore, while both the ACE and the Cognitron tasks captured deficits in memory and executive function (planning and fluency) in ALE patients, only the Cognitron tasks detected deficits in visuospatial processing and language (Supplemental Table 2). Finally, neither ACE nor Cognitron subdomains showed a deficit in the attention subdomain for patients (Supplemental Table 2). The Cognitron tasks may therefore be sensitive to cognitive changes and show improved sensitivity to a decline in visuospatial function^33^ and language abilities^7, 12^ compared to the standard neuropsychological test score.

### Memory deficits in ALE patients

#### Object Memory Task

We examined memory tasks specifically in view of ALE being a disorder associated with medial temporal lobe and hippocampal lesions^1, 7^. The Object Memory task examined immediate and delayed recall of previously seen objects. Performance was significantly above chance level in both healthy controls and ALE patients. Healthy controls outperformed the ALE patients at both immediate and delayed timepoints (main effect of group F(1, 73) = 7.40, p = .008. and time point F(1, 73) = 2.36, p = 0.13) (Figure 5A). No interaction between group and timepoint was observed (F(1,73) = 2.47, p = 0.64).

**Figure 5.**
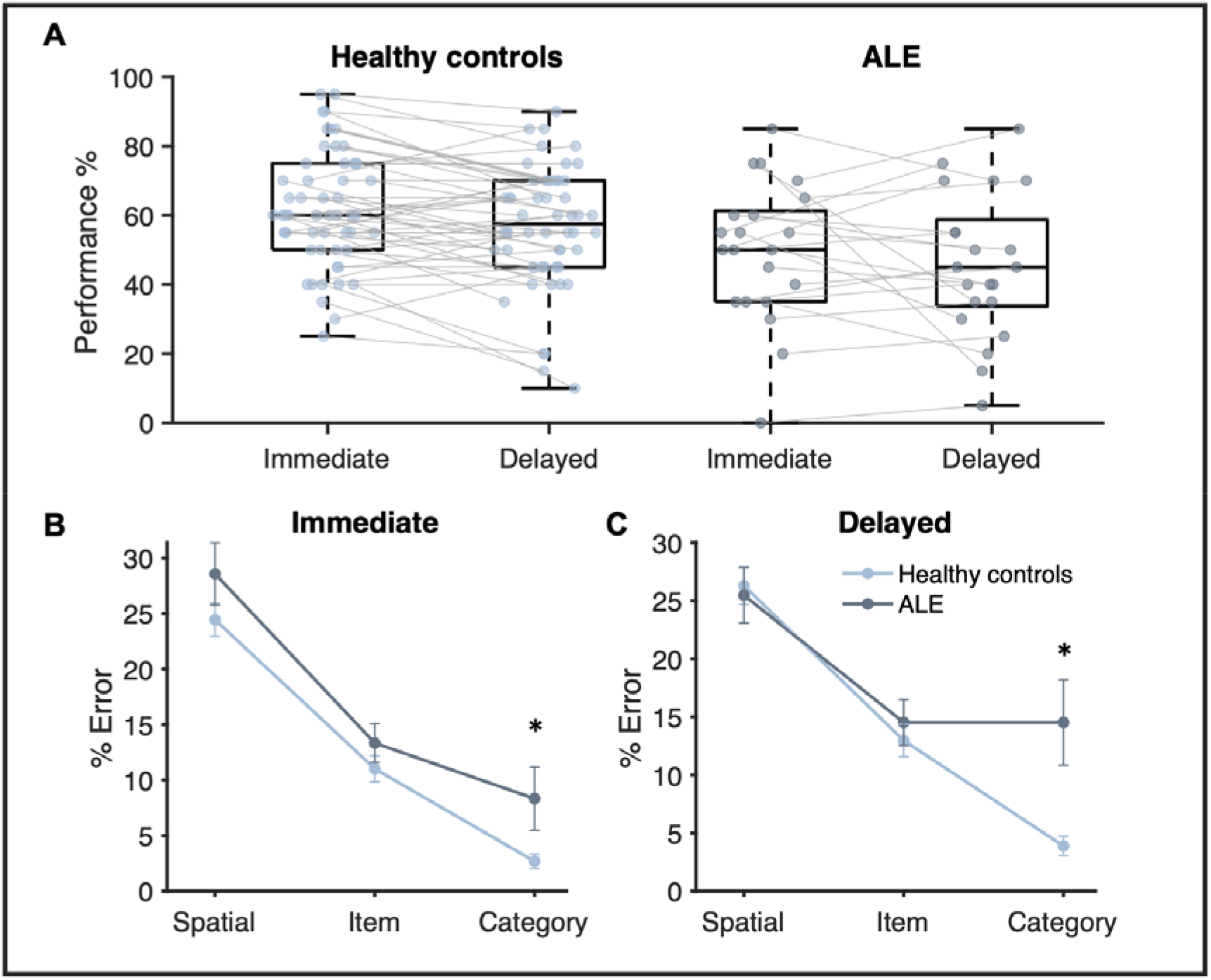
Memory performance in ALE patients compared to healthy controls. A) Performance accuracy of the object memory score by group and delay. Both groups performed above chance for both conditions. B) % Error for each type of error made for Immediate recall. Category errors were elevated in ALE compared to healthy controls. Errors bars are SEM. C) % Error for each type of error made for Delayed recall. Category errors persist at delayed recall as well.

To assess differences in performance between the two groups, we examined the types of errors made for all incorrect trials. Overall errors made on the task could be categorised into three types: 1) Spatial Error, 2) Item Error and 3) Category Error, in decreasing order of memory precision (Figure 2B). A logistic mixed effects model *[Accuracy(correct/incorrect) ∼ 1 + Timepoint + Group*ErrorType + (1+ ErrorType | Subject)]* examining the effect of group (ALE/healthy controls), error type (Spatial/Item/Category Error) and delay (Immediate or Delayed) on performance was performed. While there was no main effect of the group on overall performance (β= 0.061, 95% CI = [-1.27, 0.25], t(8993) = 0.64, p = .52), ALE patients made a significantly higher percentage of category errors compared to healthy controls (Group by category error interaction, β= 1.17, 95% CI = (0.48, 1.859), t(8993) = 3.34, p < .001. This highlights the behavioural difference in memory, which appears to be driven by an increase in low-resolution errors by ALE patients who are getting the category of the objects wrong (Figure 5B&C). As expected, a main effect of timepoint, where performance is lower at delay has been observed as well (β= 0.10, 95% CI = (0.0088, 0.20), t(8993) = 2.14, p = .032). This task, therefore, is able to identify a memory deficit in chronic ALE patients that is not detected by the spatial and digit working memory span tasks which were not significant between the groups.

### Word Memory Task

The Word Memory task had comparable immediate and delayed recall demands to the Object Memory task. In this task, ALE patients generally performed worse than healthy controls, mainly in the delayed time point (main effect of group F(1, 73) = -2.53, p = .012 and time point F(1, 73) = -2,00, p = .046). At immediate recall, no significant differences were observed in the ability to reject lures in ALE (M = 5.71, SD = 0.72) compared to healthy controls (M = 5.87, SD = 0.34), t(23.56) = 9.96, p = .35, d = 0.33), but healthy controls (M = 5.67, SD = 0.58) were more successful at rejecting foils compared to ALE patients (M = 5.10, SD = 1.04), t(25.0) = 2.37, p = .026, d = 0.77).

#### Hippocampal volume is associated with Cognitron performance

Finally, we assessed the extent of hippocampal atrophy associated with ALE. ALE patients (M = 6881.98, SD = 1010.28) compared to healthy controls (M = 7498.65, SD = 575.67) had a significantly smaller bilateral hippocampal volume (t(25.99) = 2.61, p = .0015, d = 0.84). By comparison, there was no difference in the amygdala volume which is also a region of the medial temporal lobe, between ALE patients (M = 2618.22, SD = 392.56) and healthy controls (M = 2552.92, SD = 362.05), (t(35.84) = -0.65, p = .52, d = 0.18). This indicates a specificity of atrophy to the hippocampus.

A linear regression between global Cognitron performance and total hippocampal volumes was performed across all study participants while controlling for age, gender, and years of education. This showed that a smaller hippocampal volume results in lower global cognitive scores (r(62) = 0.37, p < .001). When looking at the groups separately, the correlation was significant in the ALE patients (r(15) = 0.67, p = .017) and not in healthy controls (r(42) = 0.08, p = .42). These correlations were driven by the patients with low hippocampal volume and expected low behavioural performance. All 4 subdomains of the Cognitron correlated with hippocampus volume (all p < .025) with the strongest relationship of the hippocampus to the language domain (r = 0.28) (Supplemental Figure 3).

## Discussion

In the present study, the cognitive profile of ALE patients was compared to age-and gender-matched healthy controls using remotely administered online computerised tasks. People with ALE often present with cognitive deficits and focal atrophy of the medial temporal lobe^1, 5, 7, 34–36^. Therefore, it can serve as a model disease to study long-term cognitive impairments. Here, residual cognitive deficits in ALE (memory, language, visuospatial and planning) were found using the online platform and the outcome measures of this platform were robustly correlated with standard cognitive screening. The online Cognitron platform also had higher sensitivity compared to the ACE to detect impairments in visuospatial abilities and language (Figure 3A&B). Hippocampus-associated deficits in spatial and memory abilities were also evident, with an impairment in object memory, word memory and mental rotations tasks. Finally, hippocampal volume was correlated to cognitive metrics from this online remote assessment, where a smaller volume was associated with worse deficits. The data support the validity of remote digital testing as a clinical platform to assess long-term cognition in chronic ALE. This platform may also have the potential for longitudinal monitoring which is not possible with traditional supervised assessment methods.

ALE patients, compared to healthy controls, performed worse across most domains including memory, visuospatial performance, and higher-order executive functions such as word definitions, verbal analogies, and planning. Such differences were reported with medium to large effect sizes (Cohen’s d > 0.7). The deficits in 10 out of 14 tasks from the Cognitron battery are aligned with previous in-person studies reporting long-term cognitive deficits in ALE^9^.

The absence of deficits in other domains (attention and emotion processing) may reflect the relative specificity of the antibody-mediated brain lesion in ALE. Target detection, a measure of attention, did not differ between groups, mirroring the pen-and-paper results (Attention domain in ACE). No difference in emotional discrimination was also found between the two groups, in keeping with the limited evidence for changes in affective processes and social cognition in ALE. One previous study reported 2 out of 3 cases with deficits in fear recognition^37^. However, one patient was tested in the acute phase, which can be complicated by the neurological and psychiatric features. Together, the results indicate that emotional processing and target detection may not be affected in *chronic* ALE patients.

We did not observe differences between ALE and healthy controls in working memory span tasks, although impairment in such tasks has been described^7^ (recovery of span tasks after 3 months has also been reported^38^). Importantly, longer-term memory deficits were observed in the Object and Word Memory tasks. In the Object Memory task, we used everyday objects, available as a visual template and easily integrated into cortical regions^39, 40^ to facilitate encoding and retention of visual stimuli. A deficit in both the Object Memory and Word Memory tasks was found in immediate and delayed recall. In particular, an increase in guessing was observed in ALE during the object memory task. These results align with previous reports of memory deficits^7, 41, 42^. Cognitron tasks also revealed visuospatial processing and language deficits in ALE patients which have been reported in previous cohorts^7, 12, 43^.

Classically, the hippocampus is known to serve mnemonic and spatial functions^44^. In ALE, autoimmune antibodies target limbic structures, particularly the hippocampus^1, 5, 34–36^ so it is unsurprising to observe worse spatial and memory performance in these patients. As expected, the ALE patients studied here had hippocampal atrophy compared to healthy controls. Furthermore, hippocampal atrophy was related to the global score of cognition, including scores of memories and visuospatial capacity. This opens the door to probing hippocampal function using digital testing which will be helpful in other patient groups such as MCI and AD where the hippocampus is affected relatively early on in AD disease progression.

When comparing the Cognitron battery of tasks to standard neuropsychological tests, the cognitive composite score correlated with total ACE scores. This was particularly pronounced in the patient group. Furthermore, the subdomains of the ACE and the Cognitron tasks were highly related. The expected deficits in language and visuospatial abilities were not captured by the in-person ACE, suggesting that the computerised tasks have greater sensitivity compared to pen-and-paper tests in these domains. Although standard neuropsychological tests remain the gold standard for cognitive testing, these results present evidence for the validity of remotely administered online cognitive assessments and supports its use as a valid alternative to traditional neuropsychological testing. There are several other major benefits. Firstly, online remote assessment can allow for more widespread and convenient access to cognitive testing with high ecological validity.

Secondly, remote online testing can facilitate the collection of larger sample sizes cheaply, as has been shown in Covid-19-related cognitive studies^26, 27^. Smaller study cohorts often face the concern of selection bias that may skew cognitive profiles. Digital assessments will allow recruitment from a wide participant pool and reduce the bias of socioeconomic status, cultural biases, and accessibility to testing sites. Thirdly, long-term follow-up can be performed, which is particularly relevant when assessing cognitive decline in neurodegenerative conditions^45^ Fourthly, immediate feedback can be provided which can be informative and motivating for participants and caretakers. Finally, although not leveraged in this study, detailed time courses of task performance can be used as a basis for computational modelling approaches that improve domain precision and sensitivity of cognitive ability estimates. These advantages make digital cognitive assessment an appealing research and screening tool for neurodegenerative disorders.

Alongside molecular biomarkers, digital cognitive profiles may provide useful insight for treatment monitoring, prognostication, and improved diagnosis. We present a platform with the ability to collect self-administered cognitive data from a rare patient group with additional neuroimaging metrics. The global score of Cognitron measures correlated with in-person neuropsychological testing (ACE) and hippocampal volume, providing validity of online testing. Expected deficits not identified by the ACE were identified using Cognitron. This approach may consistently measure behavioural changes across patients and generalise to other neurological diseases with cognitive deficits, providing a novel clinical platform for neurodegenerative disease.

## Author Contribution Statement

K.S. & M.H. designed the study. K.S. collected the behavioural data. B.A collected and processed the MRI scans. K.S. analysed the data with the help of B.A., X.T., & S.G.M. W.T., P.J.H. & A.H. designed and created the online cognitive testing platform. S.R.I., S.G.M., & M.H. provided access to ALE patients. K.S. interpreted the results and drafted the manuscript with the input from B.A., X.T., A.H., S.R.I., S.G.M, and M.H. All authors read and approved the final version of the manuscript.

## Data availability statement

The datasets generated and analysed during the current study are available from the corresponding author on reasonable request.

## Ethics statements

### Patient consent for publication

Participants gave informed consent to participate in the study before taking part.

## Ethics approval

The study received approval from Oxford University’s Ethics Committee (18/SC/0048 & REC16/YH/0013).

### Open Access Statement

For the purpose of Open Access, the author has applied a CC BY public copyright licence to any Author Accepted Manuscript (AAM) version arising from this submission. The views expressed are those of the authors and not necessarily those of the NHS, the NIHR or the Department of Health.

## Data Availability

All data produced in the present study are available upon reasonable request to the authors.

## Acknowledgments

This work was funded by Wellcome Trust Principal Research Fellowship to M.H., Medical Research Council CSF [MR/P00878X] and National Institute for Health Research (NIHR) Oxford Biomedical Research Centre (BRC) to S.G.M. and in part by a senior clinical fellowship from the Medical Research Council [MR/V007173/1], Wellcome Trust Fellowship [104079/Z/14/Z], and by the NIHR Oxford BRC to S.R.I. Additional funding by the Berrow Foundation to K.S. and the Rhodes Scholarship to B.A. supported this work. The funders had no role in the study design, data collection, analysis, decision to publish, or preparation of the manuscript.

## Supplemental Material

**Supplemental Table 1.** Performance on Cognitron battery. Differences in performance in Cognitron tasks between ALE patients and healthy controls compared using Welch’s T-test. Cohen’s d is reported as a measure of effect size. The significant t-tests are highlighted in bold.

|  |  | All<br>(n = 75) | Healthy control<br>(n = 54) | ALE<br>(n = 21) | Statistics |
| --- | --- | --- | --- | --- | --- |
| 1 | Object Memory - Immediate | 47.65 (7.50) | 49.09 (5.98) | 43.95 (9.67) | **t(26.18) = 2.27, p = .031, d = 0.71** |
| 2 | Word Memory - Immediate | 21.37 (2.05) | 21.17 (1.71) | 20.14 (2.35) | **t(28.61) = 3.03, p = .0051, d = 0.90** |
| 3 | Blocks | 8.57 (2.94) | 9.24 (2.63) | 6.86 (3.05) | **t(32.23) = 3.15, p = .0040, d = 0.86** |
| 4 | Tower of London | 5.83 (2.52) | 6.30 (2.27) | 4.62 (2.78) | **t(30.91) = 2.46, p = .020, d = 0.69** |
| 5 | Digit Span | 6.31 (1.62) | 6.35 (1.46) | 6.19 (1.99) | t(28.88) = 0.34, p = .74, d = 0.10 |
| 6 | Spatial Span | 5.12 (1.08) | 5.28 (0.96) | 4.71 (1.27) | t(29.32) = 1.84, p = .076, d = 0.53 |
| 7 | Verbal Analogies | 20.43 (9.92) | 22.98 (8.59) | 13.86 (10.29) | **t(31.44) = 3.61, p = .0011, d = 1.00** |
| 8 | Target Detection | 47.47 (15.74) | 49.59 (14.11) | 42.00 (18.60) | t(29.40) = 1.69, p = .10, d = 0.49 |
| 9 | Emotional Discrimination | 42.48 (3.78) | 42.72 (3.15) | 41.86 (5.10) | t(26.15) = 0.73, p = .47, d = 0.23 |
| 10 | Word Definitions | 17.45 (3.05) | 18.43 (2.09) | 14.95 (3.71) | **t(25.10) = 4.05, p < .001, d = 1.32** |
| 11 | 2D Mental Rotation | 20.61 (6.81) | 21.87 (6.46) | 17.38 (6.75) | **t(35.11) = 2.62, p = .013, d = 0.68** |
| 12 | 3D Mental Rotation | 4.35 (6.28) | 5.89 (4.57) | 0.38 (8.21) | **t(24.97) = 2.90, p = .0076, d = 0.95** |
| 13 | Object Memory - Delayed | 45.31 (8.64) | 47.22 (6.62) | 40.38 (11.15) | **t(25.68) = 2.63, p = .014, d = 0.84** |
| 14 | Word Memory - Delayed | 19.68 (2.69) | 20.17 (2.35) | 18.43 (3.12) | **t(29.27) = 2.31, p = .028, d = 0.67** |

**Supplemental Table 2.**
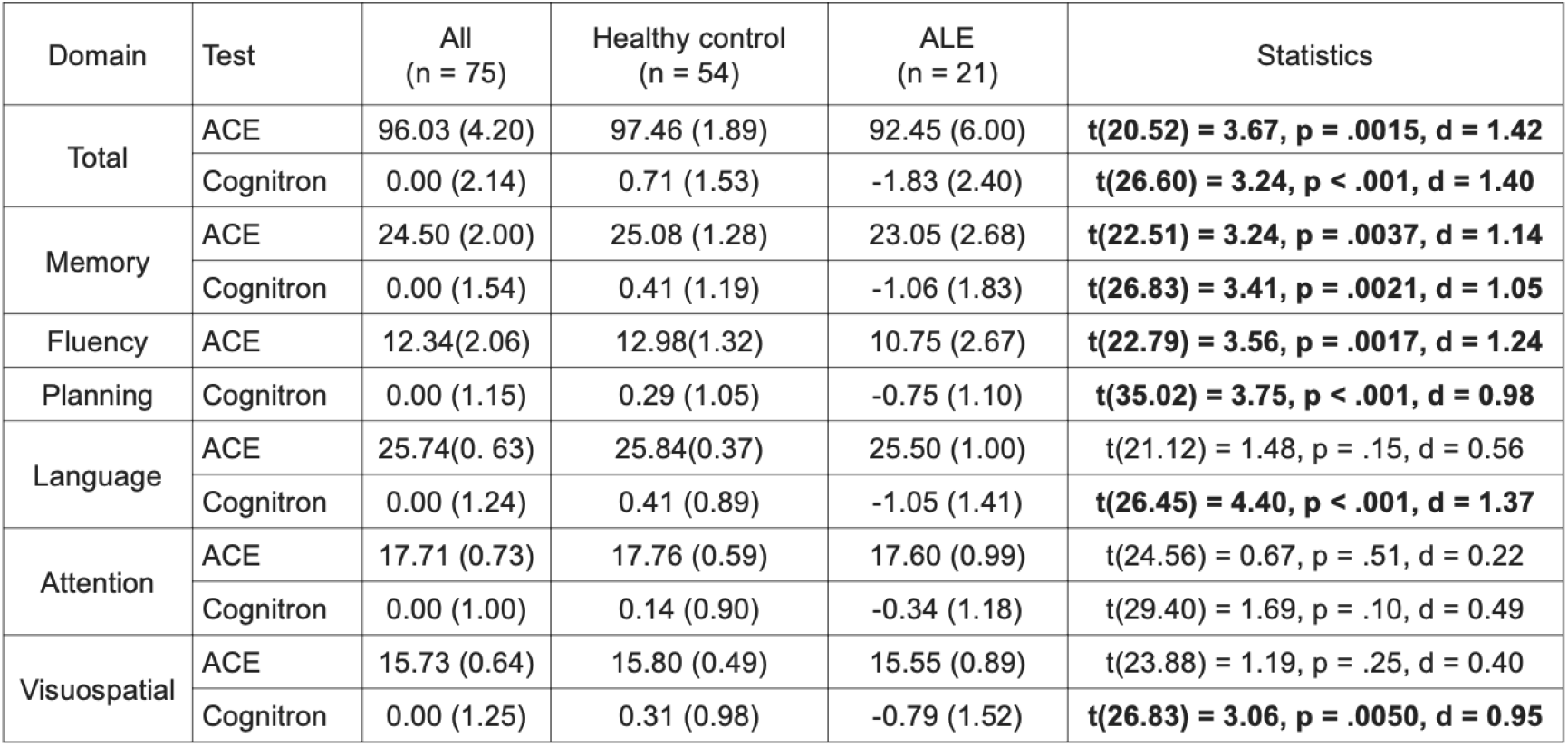
Performance on Cognitron subdomains and ACE subdomains. Differences in performance in each cognitive subdomain between ALE patients and healthy was compared using Welch’s T-test. Subdomains for the Cognitron tests were calculated by taking the first principal component of all tasks that make up the corresponding subdomain. Cohen’s d is reported as a measure of effect size. The significant t-tests are highlighted in bold.

| Domain | Test | All<br>(n = 75) | Healthy control<br>(n = 54) | ALE<br>(n = 21) | Statistics |
| --- | --- | --- | --- | --- | --- |
| Total | ACE | 96.03 (4.20) | 97.46 (1.89) | 92.45 (6.00) | **t(20.52) = 3.67, p = .0015, d = 1.42** |
|  | Cognitron | 0.00 (2.14) | 0.71 (1.53) | -1.83 (2.40) | **t(26.60) = 3.24, p < .001, d = 1.40** |
| Memory | ACE | 24.50 (2.00) | 25.08 (1.28) | 23.05 (2.68) | **t(22.51) = 3.24, p = .0037, d = 1.14** |
|  | Cognitron | 0.00 (1.54) | 0.41 (1.19) | -1.06 (1.83) | **t(26.83) = 3.41, p = .0021, d = 1.05** |
| Fluency | ACE | 12.34(2.06) | 12.98(1.32) | 10.75 (2.67) | **t(22.79) = 3.56, p = .0017, d = 1.24** |
| Planning | Cognitron | 0.00 (1.15) | 0.29 (1.05) | -0.75 (1.10) | **t(35.02) = 3.75, p < .001, d = 0.98** |
| Language | ACE | 25.74(0.63) | 25.84(0.37) | 25.50 (1.00) | t(21.12) = 1.48, p = .15, d = 0.56 |
|  | Cognitron | 0.00 (1.24) | 0.41 (0.89) | -1.05 (1.41) | **t(26.45) = 4.40, p < .001, d = 1.37** |
| Attention | ACE | 17.71 (0.73) | 17.76 (0.59) | 17.60 (0.99) | t(24.56) = 0.67, p = .51, d = 0.22 |
|  | Cognitron | 0.00 (1.00) | 0.14 (0.90) | -0.34 (1.18) | t(29.40) = 1.69, p = .10, d = 0.49 |
| Visuospatial | ACE | 15.73 (0.64) | 15.80 (0.49) | 15.55 (0.89) | t(23.88) = 1.19, p = .25, d = 0.40 |
|  | Cognitron | 0.00 (1.25) | 0.31 (0.98) | -0.79 (1.52) | **t(26.83) = 3.06, p = .0050, d = 0.95** |

**Supplemental Figure 1.**
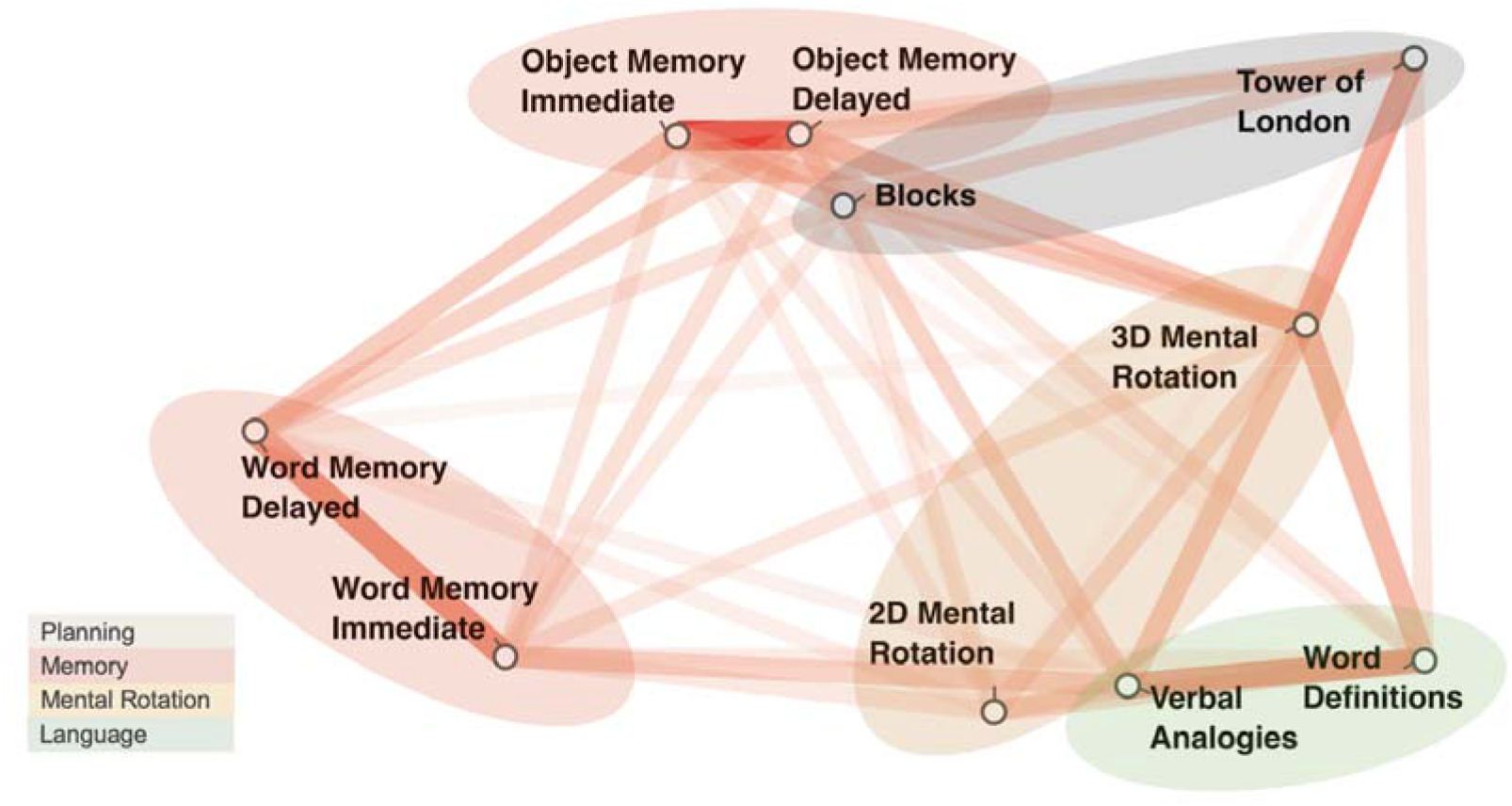
Cognitron task correlation. Network plot of task correlations (healthy controls and ALE patients combined, n = 75). Only metrics that significantly correlated with another task are included in the plot. A smaller distance and lower opacity of the connecting line indicate a stronger correlation.

**Supplemental Figure 2.**
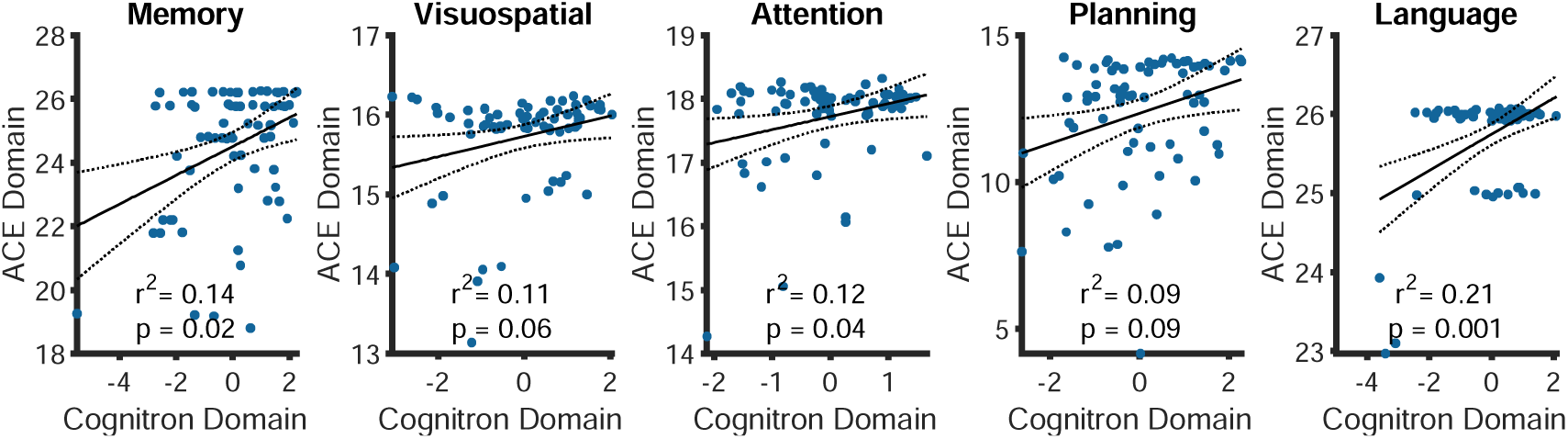
Correlation between ACE and Cognitron subdomains. Correlation between clinical ACE score subdomains and digital Cognitron score subdomains. Each domain from the neuropsychological test correlates with the Cognitron scores.

**Supplemental Figure 3.**
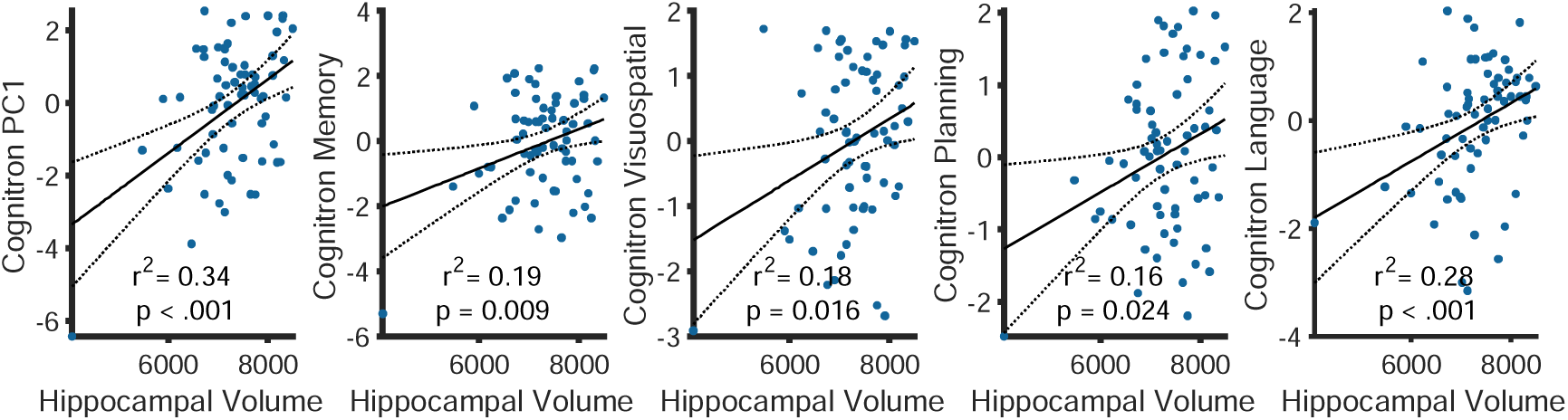
Global Cognitron score and hippocampal volume. Correlation between global Cognitron score and bilateral hippocampal volume.

## References

1. Irani, S. R. et al. Antibodies to Kv1 potassium channel-complex proteins leucine-rich, glioma inactivated 1 protein and contactin-associated protein-2 in limbic encephalitis, Morvan’s syndrome and acquired neuromyotonia. Brain 133, 2734–2748 (2010).

2. Ramberger, M. et al. Distinctive binding properties of human monoclonal LGI1 autoantibodies determine pathogenic mechanisms. Brain 143, 1731–1745 (2020).

3. Joubert, B. et al. Human CASPR2 Antibodies Reversibly Alter Memory and the CASPR2 Protein Complex. Ann. Neurol. 91, 801–813 (2022).

4. Thompson, J. et al. The importance of early immunotherapy in patients with faciobrachial dystonic seizures. Brain 141, 348–356 (2018).

5. Irani, S. R. et al. Faciobrachial dystonic seizures: the influence of immunotherapy on seizure control and prevention of cognitive impairment in a broadening phenotype. Brain 136, 3151–3162 (2013).

6. Griffith, S. P., Malpas, C. B., Alpitsis, R., O’Brien, T. J. & Monif, M. The neuropsychological spectrum of anti-LGI1 antibody mediated autoimmune encephalitis. J. Neuroimmunol. 345, 577271 (2020).

7. Finke, C. et al. Evaluation of Cognitive Deficits and Structural Hippocampal Damage in Encephalitis With Leucine-Rich, Glioma-Inactivated 1 Antibodies. JAMA Neurol. 74, 50 (2017).

8. Nantes, J. C. et al. Hippocampal Functional Dynamics Are Clinically Implicated in Autoimmune Encephalitis With Faciobrachial Dystonic Seizures. Front. Neurol. 9, 736 (2018).

9. Binks, S. N. M. et al. Residual Fatigue and Cognitive Deficits in Patients After Leucine-Rich Glioma-Inactivated 1 Antibody Encephalitis. JAMA Neurol. 78, 617 (2021).

10. Titulaer, M. J. et al. Treatment and prognostic factors for long-term outcome in patients with anti-NMDA receptor encephalitis: an observational cohort study. Lancet Neurol. 12, 157–165 (2013).

11. Rodriguez, A. et al. LGI1 antibody encephalitis: acute treatment comparisons and outcome. J. Neurol. Neurosurg. Psychiatry 93, 309–315 (2022).

12. Bettcher, B. M. et al. More than memory impairment in voltage-gated potassium channel complex encephalopathy. Eur. J. Neurol. 21, 1301–1310 (2014).

13. Hang, H.-L., Zhang, J.-H., Chen, D.-W., Lu, J. & Shi, J.-P. Clinical Characteristics of Cognitive Impairment and 1-Year Outcome in Patients With Anti-LGI1 Antibody Encephalitis. Front. Neurol. 11, 852 (2020).

14. Day, G. S. Rethinking Outcomes in Leucine-Rich, Glioma-Inactivated 1 Protein Encephalitis: “Good” Isn’t Good Enough. JAMA Neurol. 74, 19 (2017).

15. Gadoth, A. et al. Expanded phenotypes and outcomes among 256 LGI1/CASPR2-IgG-positive patients: LGI1/CASPR2-IgG ^+^ Patients. Ann. Neurol. 82, 79–92 (2017).

16. van Sonderen, A. et al. Anti-LGI1 encephalitis: Clinical syndrome and long-term follow-up. Neurology 87, 1449–1456 (2016).

17. Frisch, C., Malter, M. P., Elger, C. E. & Helmstaedter, C. Neuropsychological course of voltage-gated potassium channel and glutamic acid decarboxylase antibody related limbic encephalitis. Eur. J. Neurol. 20, 1297–1304 (2013).

18. Heine, J. et al. Beyond the limbic system: disruption and functional compensation of large-scale brain networks in patients with anti-LGI1 encephalitis. J. Neurol. Neurosurg. Psychiatry 89, 1191–1199 (2018).

19. Pertzov, Y. et al. Binding deficits in memory following medial temporal lobe damage in patients with voltage-gated potassium channel complex antibody-associated limbic encephalitis. Brain 136, 2474–2485 (2013).

20. Ding, Z., Lee, T. & Chan, A. S. Digital Cognitive Biomarker for Mild Cognitive Impairments and Dementia: A Systematic Review. J. Clin. Med. 11, 4191 (2022).

21. Öhman, F., Hassenstab, J., Berron, D., Schöll, M. & Papp, K. V. Current advances in digital cognitive assessment for preclinical Alzheimer’s disease. Alzheimers Dement. Diagn. Assess. Dis. Monit. 13, (2021).

22. Doraiswamy, P. M. et al. Validity of the Web-Based, Self-Directed, NeuroCognitive Performance Test in Mild Cognitive Impairment. J. Alzheimers Dis. 86, 1131–1136 (2022).

23. Del Giovane, M. et al. Computerised cognitive assessment in patients with traumatic brain injury: an observational study of feasibility and sensitivity relative to established clinical scales. eClinicalMedicine 59, 101980 (2023).

24. Brooker, H. et al. FLAME: A computerized neuropsychological composite for trials in early dementia. Alzheimers Dement. Diagn. Assess. Dis. Monit. 12, (2020).

25. Giunchiglia, V., et al. Iterative decomposition of visuomotor, device and cognitive variance in large scale online cognitive test data. https://www.researchsquare.com/article/rs-2972434/v1<x> (2023) doi:10.21203/rs.3.rs-2972434/v1.

26. Hampshire, A. et al. Cognitive deficits in people who have recovered from COVID-19. EClinicalMedicine 39, 101044 (2021).

27. Hampshire, A. et al. Multivariate profile and acute-phase correlates of cognitive deficits in a COVID-19 hospitalised cohort. eClinicalMedicine 47, 101417 (2022).

28. Zhao, S. et al. Rapid vigilance and episodic memory decrements in COVID-19 survivors. Brain Commun. 4, fcab295 (2022).

29. Mueller, C. et al. Review and meta-analysis of neuropsychological findings in autoimmune limbic encephalitis with autoantibodies against LGI1, CASPR2, and GAD65 and their response to immunotherapy. Clin. Neurol. Neurosurg. 224, 107559 (2023).

30. Hsieh, S., Schubert, S., Hoon, C., Mioshi, E. & Hodges, J. R. Validation of the Addenbrooke’s Cognitive Examination III in Frontotemporal Dementia and Alzheimer’s Disease. Dement. Geriatr. Cogn. Disord. 36, 242–250 (2013).

31. Alfaro-Almagro, F. et al. Image processing and Quality Control for the first 10,000 brain imaging datasets from UK Biobank. NeuroImage 166, 400–424 (2018).

32. Voevodskaya, O. et al. The effects of intracranial volume adjustment approaches on multiple regional MRI volumes in healthy aging and Alzheimer’s disease. Front. Aging Neurosci. 6, (2014).

33. Galioto, R., Aboseif, A., Krishnan, K., Lace, J. & Kunchok, A. Cognitive outcomes in anti-LGI-1 encephalitis. J. Int. Neuropsychol. Soc. 29, 541–550 (2023).

34. Bien, C. G. et al. Immunopathology of autoantibody-associated encephalitides: clues for pathogenesis. Brain 135, 1622–1638 (2012).

35. Bien, C. G. et al. Limbic encephalitis as a precipitating event in adult-onset temporal lobe epilepsy. Neurology 69, 1236–1244 (2007).

36. Irani, S. R. et al. Faciobrachial dystonic seizures precede Lgi1 antibody limbic encephalitis. Ann. Neurol. 69, 892–900 (2011).

37. Dodich, A. et al. Neuropsychological and FDG-PET profiles in VGKC autoimmune limbic encephalitis. Brain Cogn. 108, 81–87 (2016).

38. Krastinova, E., Vigneron, M., Le Bras, P., Gasnault, J. & Goujard, C. Treatment of limbic encephalitis with anti-glioma-inactivated 1 (LGI1) antibodies. J. Clin. Neurosci. 19, 1580– 1582 (2012).

39. Wang, S.-H. & Morris, R. G. M. Hippocampal-Neocortical Interactions in Memory Formation, Consolidation, and Reconsolidation. Annu. Rev. Psychol. 61, 49–79 (2010).

40. Dudai, Y., Karni, A. & Born, J. The Consolidation and Transformation of Memory. Neuron 88, 20–32 (2015).

41. Miller, T. D. et al. Focal CA3 hippocampal subfield atrophy following LGI1 VGKC-complex antibody limbic encephalitis. Brain 140, 1212–1219 (2017).

42. Hanert, A. et al. Hippocampal Dentate Gyrus Atrophy Predicts Pattern Separation Impairment in Patients with LGI1 Encephalitis. Neuroscience 400, 120–131 (2019).

43. Galioto, R., Aboseif, A., Krishnan, K., Lace, J., & Kunchok, A. Cognitive outcomes in anti-LGI-1 encephalitis. J. Int. Neuropsychol. Soc.

44. Olton, D. S., Becker, J. T. & Handelmann, G. E. Hippocampus, space, and memory. Behav. Brain Sci. 2, 313–322 (1979).

45. Wesnes, K. A. et al. Utility, reliability, sensitivity and validity of an online test system designed to monitor changes in cognitive function in clinical trials: Online cognitive testing in clinical trials. Int. J. Geriatr. Psychiatry 32, e83–e92 (2017).

